# SURGICALLY IMPORTANT ANATOMIC VARIATIONS OF EXTRAHEPATIC BILIARY SYSTEM AMONG BLACK KENYAN CADAVERS AT MOI UNIVERSITY HUMAN ANATOMY LABORATORY

**DOI:** 10.1101/2023.01.28.23285136

**Authors:** Dan Ndiwa, Andrew Wandera, Anthony Njoroge, Gerald Lwande, Mohamed El-Badawi

## Abstract

**Background:** Anatomical variations of the human body including the extra hepatic biliary system exist across various individuals. Understanding the variant anatomy of the extrahepatic biliary system aids surgeons in avoiding iatrogenic injuries. This is important in resource limited settings where it is not possible to perform adequate radiological investigations of the hepatobiliary system prior to surgery. This study described the anatomic variation of the extrahepatic biliary system among Kenyans.

**Methods:** This was a cross-sectional study conducted at Moi University’s Anatomy Laboratories among 42 adult cadaveric specimens. Specimen dissections were conducted as per the fifteenth edition of Cunningham’s manual of Practical Anatomy. The variant anatomy data collected were filled in a structured data collection form, analysed and presented using descriptive statistics.

**Study Findings:** Of the 42 cadavers sampled, 62% (n=26) were male while 38% (n=16) were female. All had a gall bladder being drained by the cystic duct. The length of the cystic duct ranged between 7-35 mm joining the common hepatic duct to form the common bile duct in 98% (n=41) of all the cadavers sampled. This confluence was to the left in 7.1% (n=3), right 42.9% (n=18), anteriorly in 14.3% (n=6) and posteriorly 35.7% (n=14). A single cadaver (2%) had the cystic duct drain into the right hepatic duct. Two thirds (66.7%; n=28) of the cadavers sampled had the confluence of the right and the left hepatic duct outside the liver. There were no cholecystohepatic ducts in this study.

**Conclusion:** The study determined the existence of surgically important variant anatomy of the extrahepatic biliary system. There is need for greater appreciation of the extrahepatic biliary system variant anatomy by both surgeons and radiologists so as to decrease morbidity and improve on surgical outcomes.

## Introduction

Anatomical variations within the human body are common (1) with the extrahepatic biliary system being no exception (2). Understanding the normal and variant anatomy of the extrahepatic biliary system and its blood supply when present should aid the surgeon in avoiding iatrogenic complications (3,4). These surgical procedures include hepatic lobectomy, liver transplant, pancreatic duodenoscopy, laparoscopy and other minimally invasive surgeries, cholecystectomy and vascular surgery of the liver (5). Sound knowledge of the normal and variant anatomy forms the basis for the interpretation of every imaging examination for the diagnostic and interventional radiologists (6,7).

The normal and variant anatomy can be identified through high quality imaging modalities which are not within reach and inaccessible to most patients within our resource limited setup (8,9). There is limited local data on surgically important variant anatomy of the extrahepatic biliary system and its blood supply (4,10). This study determined the variant anatomy of the extrahepatic biliary system. Knowledge of the variant anatomy is therefore important not only for biliary and minimally invasive surgeons but also radiologist, interventional radiologists and gastroenterologists.

## Materials and Methods

This was a cross sectional descriptive study involving observation of the extrahepatic biliary system anatomic patterns of cadavers seen at human anatomy laboratory of Moi University School of Medicine. The study included all the male and female cadaveric specimen at the Moi university school of medicine human anatomy laboratory according to Anatomy Act of Kenya (11). It excluded mutilated or decomposed cadavers whose anatomy on the abdominal region has been distorted. Dissection was done according to the Cunningham manual of Practical Anatomy (12). Midline incision from xiphisternum towards umbilicus. Incision extended laterally, from xiphisternum along the coastal margin. Rectus muscle cut open transversely from the umbilicus entering the abdominal cavity.

Stomach identified and its curvatures were defined. Pulling the lesser curvature, lesser omentum identified, and its right free margin was defined, and then hepatoduodenal ligament was identified. Now the greater omentum was cut transversely below it was pushed forwards towards the right. Loops of small intestine were pushed towards left and second part of duodenum was exposed. Now, the stomach was reflected fully upwards to expose the pancreas and then it was cut at the neck of the pancreas making the visceral surface of the liver, free along with duodenum and head of the pancreas. The ribs were cut open along the midaxillary line on both sides and reflected upwards along with sternum, to make the parietal surface of liver free.

Dissection of the celiac trunk to the origin and its terminal branches, common hepatic artery and its branches, superior mesenteric artery and its branches, cystic artery, the gallbladder, cystic duct, right, left and common hepatic ducts and common bile duct till it entered the second part of the duodenum were dissected in all specimens.

A structured data collection form was used to collect the data. The completed data collection form was stored in locked cabinets with restricted access. The data was entered and analysed using the SPSS version 24. Categorical variables were summarized as frequencies and the corresponding percentages. Continuous variables were presented as interquartile ranges. In order to protect and respect the rights of the participants who took part in the study. Ethical approval was obtained from the Institutional Research and Ethics Committee (IREC) of Moi University school of Medicine.

## Results

The study enrolled 42 cadavers of which 62% (n=26) were male while 38% (n=16) were female.

### Gallbladder

All the study subjects had a gall bladder sited on the right side of the falciform ligament and being drained by the cystic duct.

### Long cystic duct with low fusion with the common hepatic duct

The cystic duct length ranged from 7mm to 35 mm with a median value of 17mm. 98% (n=41) of the study subjects had cystic duct join the common hepatic duct to form the common bile duct. The cystic duct joined the common hepatic duct to form the common bile duct either to the Left (7.5%), the right (45%), anteriorly (15%) and posteriorly (32.5%) in a straight (55%), tortuous (27.5%) and spiral (17.5%) course. The common bile duct was formed by the confluence of the cystic duct and common hepatic duct in nearly all of the cadavers (95.2%; n=40).

**Table 1:** Common Bile Duct as a confluence of Cystic Duct and Common Hepatic Duct.

|  |  | Percent (n) |
| --- | --- | --- |
| **CBD formed by the confluence of CD and CHD** | Yes | 97.6 (41) |
|  | No | 2.4 (1) |
|  | Total (N) | 100 (42) |

**Image 1:**
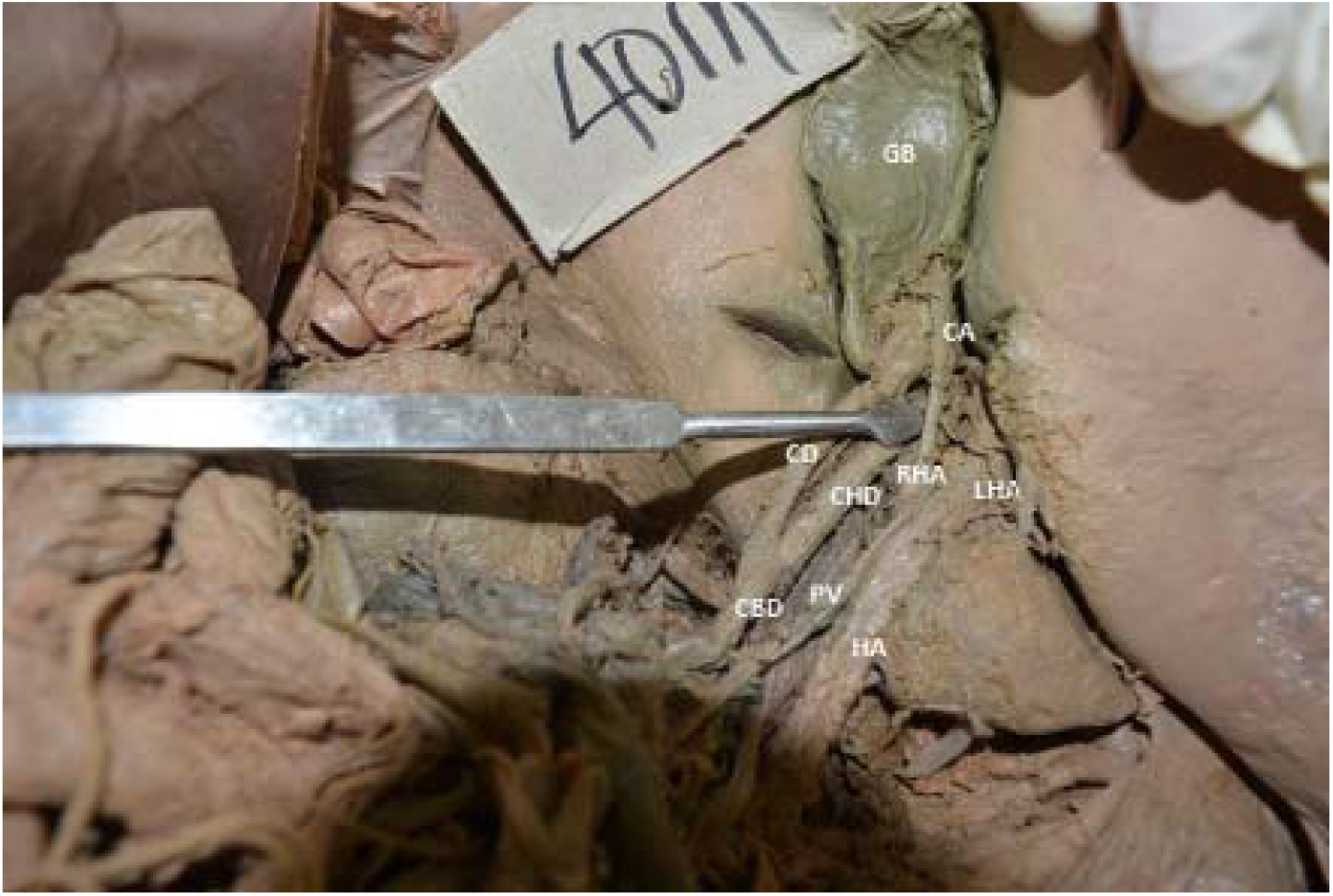
Low fusion of cystic duct with common hepatic duct. *(Key: HA: hepatic artery, LHA: left hepatic artery, RHA: right hepatic artery, CA: cystic artery, CD: cystic duct, CHD: common hepatic duct, CBD: common bile duct, PV: portal vein, GB: gall bladder)*

### Abnormal fusion of the right and left hepatic ducts with the cystic duct entering the confluence

The study observed that 33.3% (n=14) had the confluence of the right and the left hepatic duct within the liver while 66.7% (n=28) were outside the liver. However, there was no abnormal fusion of the right and left hepatic ducts with the cystic ducts entering the confluence. There was no trifurcation observed in this study.

**Table 2:** Confluence of the right and left hepatic ducts.

|  |  | Percentage (n) |
| --- | --- | --- |
| Confluence of the right and left hepatic ducts. | Within the liver | 33.3 (14) |
|  | Outside the liver | 66.7 (28) |
|  | N | 42 |

The study determined the length of the common hepatic duct to be between 1.0-4.2 cm with a mean length of 2.26 cm. There were no accessory hepatic, cholecystohepatic and accessory ducts found in this study.

### Cystic duct entering the right hepatic duct

A single cadaver (2%) had the cystic duct draining into the right hepatic duct (Image 2).

**Image 2:**
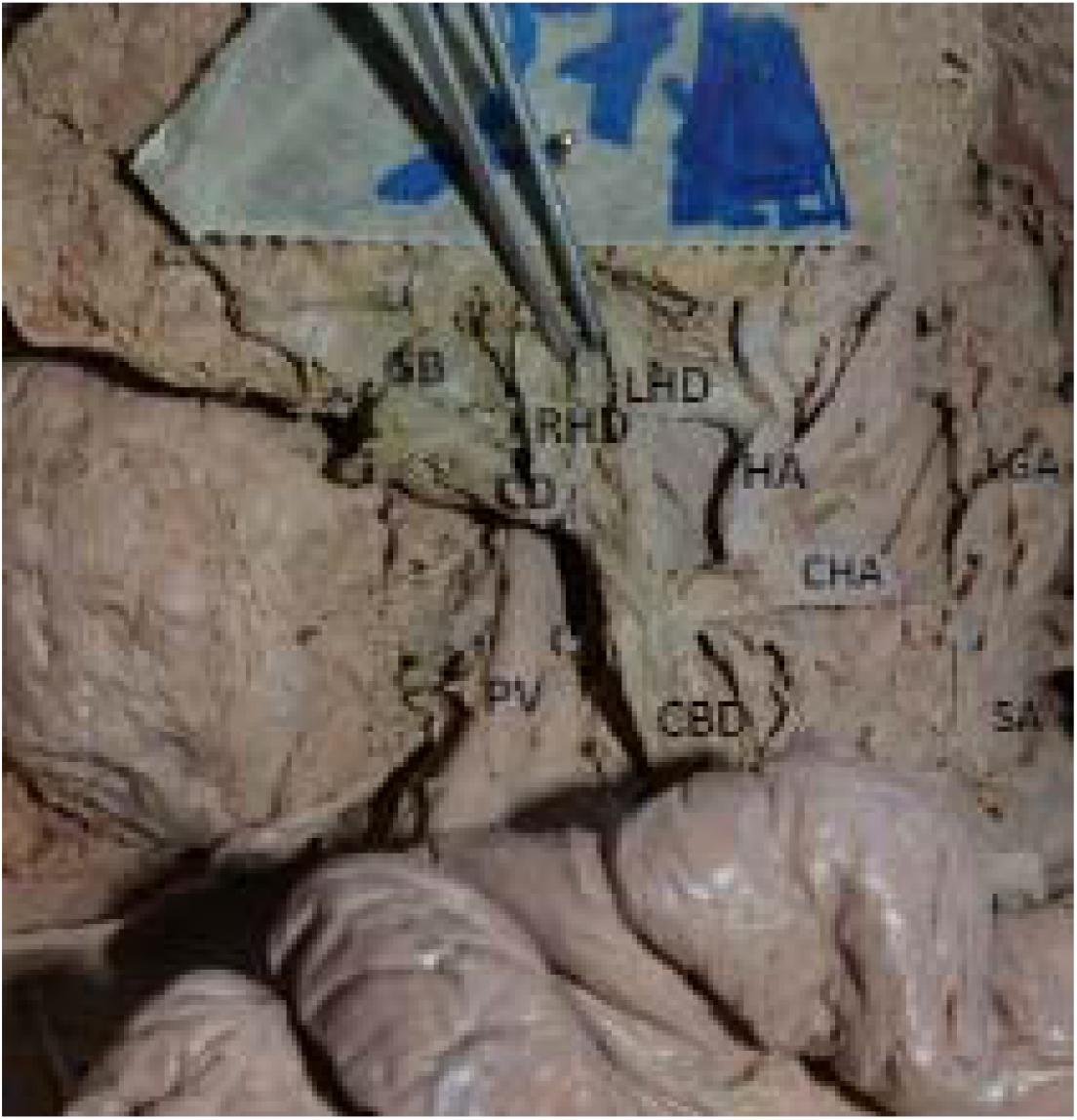
Cystic duct joining the right hepatic duct. *(Key: CHA: common hepatic artery, HA: hepatic artery, LHD: Left hepatic duct, RHD: right hepatic duct CD: cystic duct, CBD: common bile duct, PV: portal vein, GB: gall bladder)*

## Discussion

The five surgically important ductal anomalies are: Long cystic duct with low fusion with the common hepatic duct; Abnormal fusion of the right and left hepatic ducts with the cystic duct entering the confluence; Accessory hepatic ducts; Cystic duct entering the right hepatic duct and Cholecystohepatic ducts (13,14).

### Cystic Ducts

This study determined that the cystic duct’s length ranged from 0.7cm to 3.5 cm with a median value of 1.7cm. The cystic duct usually measures 2–4 cm in length (15)This variance could be attributed to the difference in the investigation technique adopted by Turner and Fulcher (2001) who used magnetic resonance imaging (15)

The study also showed that cystic duct joined the common hepatic duct to form the common bile duct either to the left (7.5%), the right (45%), anteriorly (15%) and posteriorly (32.5%) in a straight (55%), tortuous (27.5%) and spiral (17.5%) course. These findings compare to an American study which found that the cystic duct enters the extrahepatic bile duct from the right lateral aspect in 49.9% of cases, from the medial aspect (left) in 18.4%, and from an anterior or posterior position in 31.7% (16). An Indian study found a medial (left) insertion in 10%-17% (17). A single (2%) study participant had the cystic duct drained through the right hepatic duct. This corresponds to the findings of previous studies who found that cystic duct may join the right hepatic duct in 0.6%-2.3% cases (17,18).

### Hepatic Ducts

First, variations on the hepatic ducts occur at different levels of confluence of the two (right and left) hepatic ducts. The study determined that 33.3% (n=14) had the confluence of the right and the left hepatic duct within the liver while 66.7% (n=28) were outside the liver. When the confluence what outside the liver, it only ran parallel to join the cystic duct to form the common bile duct in one cadaver (3.6%). This finding is similar to a radiological study found the cystic duct ran a parallel course to the common hepatic duct in 1.5%-25% (19). However, this study like that of Cachoeira did not find accessory and Cholecystohepatic ducts (20). Cholecystohepatic ducts were not found in this study due the fact that most delicate structures collapse in formalin preserved cadavers. The study determined the length of the common hepatic duct to be between 1.0-4.2 cm (mean: 2.26 cm) which compares closely to a Brazilian study which found the length to vary between 0.4 and 5.6 cm, with a mean average of 2.17 cm (20).

## Conclusion

The study determined the existence of surgically important variant anatomy of the extrahepatic biliary system and its blood supply among black Kenyans. A small (2%) proportion of the participants had the cystic duct draining through the right hepatic duct. When the confluence of the right and left hepatic ducts was outside the liver, it ran parallel to the cystic duct to form the common bile duct. There was no marked difference in the length of the common bile duct noted between the male and female cadavers. There is need for greater appreciation of the extrahepatic biliary system’s variant anatomy and its blood supply by both surgeons and radiologists so as to decrease morbidity and improve on surgical outcomes.

## Data Availability

All data produced in the present study are available upon reasonable request to the authors

